# Detecting imported malaria infections in endemic settings using molecular surveillance: current state and challenges

**DOI:** 10.1101/2024.09.09.24313136

**Authors:** Mahdi Safarpour, Luis Cabrera Sosa, Dionicia Gamboa, Jean-Pierre Van geertruyden, Christopher Delgado-Ratto

**Affiliations:** Malaria Research Group (MaRCH), Family Medicine and Population Health Department, Global Health Institute, Faculty of Medicine and Health Sciences, University of Antwerp, Belgium; Laboratorio de Malaria: Parásitos y Vectores, Laboratorios de Investigación y Desarrollo, Facultad de Ciencias e Ingeniería, Universidad Peruana Cayetano Heredia, Lima, Peru; Instituto de Medicina Tropical “Alexander von Humboldt”, Universidad Peruana Cayetano Heredia, Lima, Peru

**Keywords:** malaria elimination, metapopulation, population genetics, travel-acquired malaria, genomic surveillance

## Abstract

The Global Technical Strategy for Malaria 2016–2030 targets eliminating malaria from at least 35 countries and reducing case incidence by 90% globally. The importation of parasites due to human mobilization presents a significant challenge to achieve elimination as it can undermine local interventions. A thorough understanding of importation is necessary to support efforts to control and further lead to elimination. Parasite genetic data is extensively deployed to investigate the space-time spread of imported infections. In this matter, this systematic review aimed to aggregate evidence on the use of parasite genetic data for mapping imported malaria and the statistical analytical methods. We discuss the advantages and limitations of the deployed genetic approaches and propose a suitable type of genetic data and statistical framework to discriminate imported malaria infections from local infections. The findings provide actionable insights for national control programs, helping them select the most suitable methods for detecting imported cases while supporting the evaluation of elimination program performance, particularly in low transmission settings.

## 1 Introduction

Over the last decade, the technical framework for malaria control and elimination recommended by the World Health Organization (WHO) has substantially reduced the global malaria burden (Feachem et al. 2019). Despite all efforts, the number of endemic countries with fewer than 10,000 malaria cases per year increased from 26 in 2000 to 47 in 2020 (Rosenthal 2022). Likewise, malaria cases increased from 227 million in 2019 to 249 million in 2022 (Venkatesan 2024). The relative increase in the number of malaria cases is favored by malaria parasite importation due to human mobility from one malaria endemic region to the other (Chang, Wesolowski, Sinha, Jacob, Mahmud, Uddin, Zaman, Hossain, Faiz, and Ghose 2019). From a fundamental perspective, this scenario reflects the metapopulation dynamics theory, where the parasite population’s subdivisions are geographically dispersed but with limited interaction between the components. Related to metapopulation dynamics, human mobility facilitates parasite importation and recolonization of areas where the parasite population was (nearly) extinct due to successful interventions (Ariey et al. 2003). Therefore, it is essential to quantify human mobility’s role in the malaria parasite’s spatial distribution and connectivity in countries aiming for malaria elimination (Tam et al. 2021).

To unravel how human mobility influences malaria’s geographical spread, researchers often collect data on recent travel history in clinical cases (Wesolowski et al. 2012). In theory, this would help to spot regions where the disease is imported from and design targeted interventions. However, the quality of travel survey data depends on the respondents’ ability to recall their travel history accurately (Wesolowski et al. 2014). Therefore, the existence of recall bias in travel survey data can limit their application to correctly infer the parasite’s origin. In contrast, mobile phone data and Global Positioning System (GPS) tracking can provide large volumes of data to measure the spatial spread of malaria parasites (Carrasco-Escobar et al. 2020). Nevertheless, the application of mobile phone data is limited to the locations where cell towers exist (Buckee et al. 2013). In addition, GPS tracking requires devices comfortable enough to carry for long periods, replace or recharge batteries quite frequently, and relies upon receiving high-quality signals from satellites and harsh conditions-resistant materials, among other logistics.

Current efforts involve the use of parasite genetic data to address the problem of determining the origin of malaria infections. In principle, the genetic information of parasites can potentially provide the most direct measure of parasite connectivity and can lead to estimating the sources and sinks of malaria parasites (Wesolowski et al. 2018). Multiple molecular techniques can be used to profile the parasite’s genetic data. Microsatellite genotyping is among the most common molecular methods to study malaria parasite genetic diversity and population structure in the Peruvian Amazon and West Africa (Manrique et al. 2019; Mobegi et al. 2012). Other methods, such as single nucleotide polymorphisms (SNPs) profiling, are widely used to explore parasite population structure in Papua New Guinea (Fola et al. 2018)and at the Myanmar-China border (Lo, Lam, et al. 2017). In recent years, with the rapid development of sequencing technologies, the whole-genome sequencing (WGS) of malaria parasites has been used as an alternative approach to study the spatial distribution of malaria parasites not only across the continents but also within local communities with smaller spatial scales such as Southeast Asia (Wasakul et al. 2023).

Despite the widespread application of different molecular techniques in malaria research, there remains a lack of information regarding their effectiveness in accurately detecting imported cases. This gap is particularly noted at smaller geographical scales, such as within countries where parasites are expected to be highly related. Consequently, this systematic review describes the current evidence, including the types of genetic data used and the methods employed to analyze the data. The findings can provide comprehensive evidence for epidemiologists and other health scientists on the reliability of malaria parasite genetic data to map the imported individuals and provide actionable insights for policymakers to eliminate malaria.

## 2 Materials and Methods

### 2.1 Search strategy

Four online bibliographic databases were searched for this review, including PubMed, Scopus, Cochrane Library, and Web of Science. We developed a comprehensive search strategy based on the keywords related to malaria parasite species, human mobility, and molecular/genomic tools used to map the spatial spread of malaria infection (Details are provided in Appendix 1). This review was restricted to studies reported in English without considering any time limitation. Full details of the search strategy are available in supplementary material. Furthermore, reference lists of included articles were screened to identify eligible studies not found through our initial search.

### 2.2 Study selection

Articles from online bibliographic databases were stored and combined as EndNote files, where duplicate articles were removed. The titles and abstracts of articles were screened, and irrelevant articles were excluded. Articles selected for full-text review were screened again for final decision of inclusion. Two reviewers screened full texts independently, and disagreements were resolved by consensus. Studies were included if they were peer-reviewed articles, used parasite genetic data, studied either *Plasmodium vivax* or *Plasmodium falciparum*, and contained a statistical method that investigated the origin of infection or explicitly accounted for connections between geographical areas either at a global level or local level. Articles were excluded if they only descriptively reported the genetic data without integrating them into a statistical method. They were case reports or case series studies, conference abstracts, meeting reports, workshop proceedings, or review articles.

### 2.3 Data extraction

Data extraction was done from the full text of all eligible articles. Data related to study setting, species of malaria parasites, type of infection, sample size, and type of genetic markers were extracted. Furthermore, the available data on drug resistance markers, travel surveys, and GPS tracking were recorded for each study. The statistical methods used for analyzing the parasite genetic data were also collected.

### 2.4 Data synthesis

Data were synthesized narratively, and descriptive statistics were used to summarize the main study characteristics, such as study setting, sample size, and the number of genetic loci examined. Studies were grouped according to the following three criteria: (1) species of malaria parasites (*Plasmodium vivax* or *Plasmodium falciparum*); (2) type of genetic marker (SNP barcodes, Microsatellite, or Whole-genome sequencing) and (3) statistical approach. Findings were descriptively presented and discussed using the genetic marker type used for genotyping and the statistical approach used to estimate the origin of infection.

## 3 Results

A total of 584 articles were initially retrieved from the databases, resulting in 479 unique publications after removing duplicates. We first screened the titles and abstracts, and 129 articles were promoted for full-text review. We could identify one additional article by manually searching the reference lists of included articles. Of those, 96 studies were excluded because they did not meet the inclusion criteria. The full-text review resulted in identifying 33 studies that were eligible for inclusion. The PRISMA flow chart illustrating the screening process is summarized in Figure 1.

**Figure 1.**
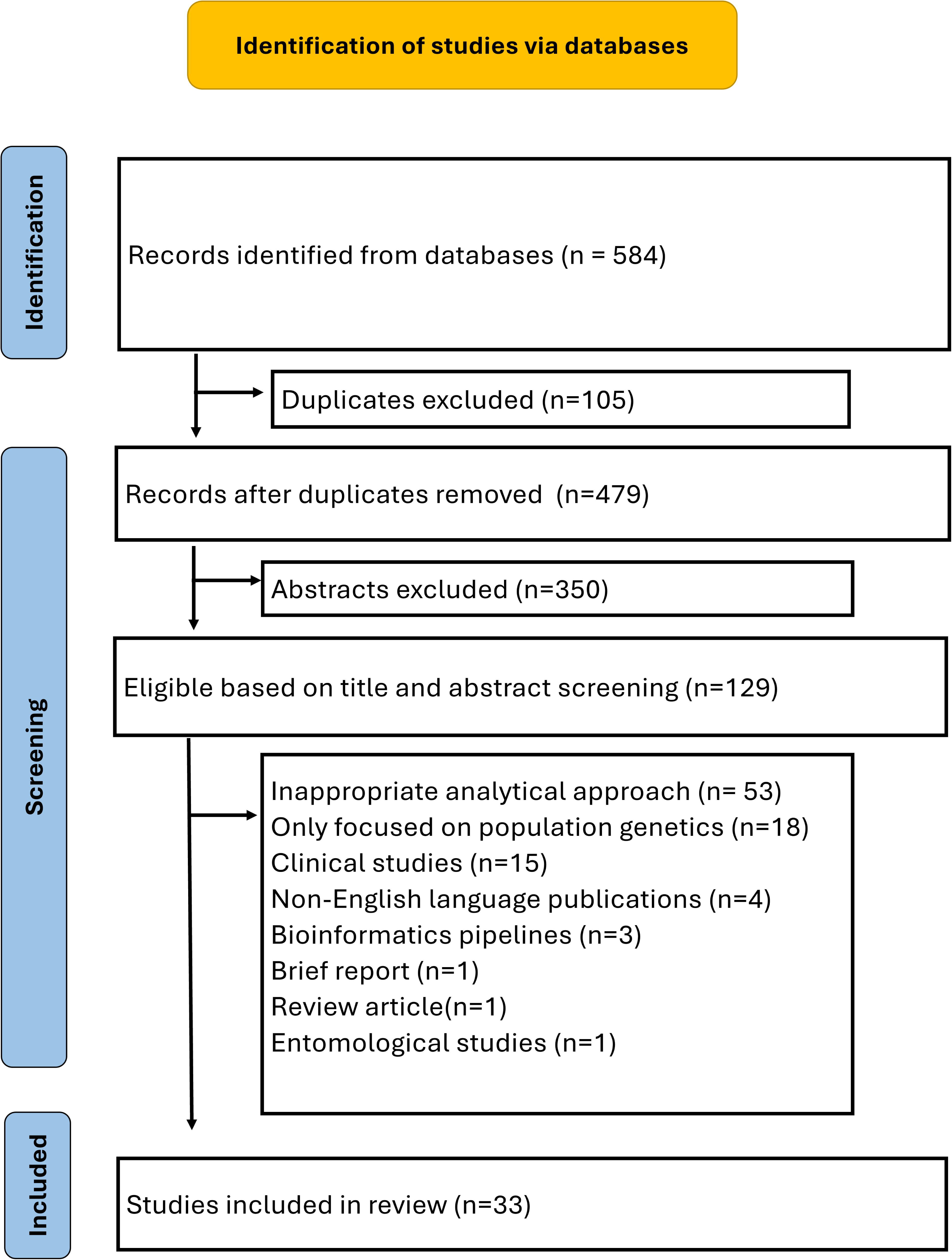
The timeline of eligible studies published across each continent over time

### 3.1 General characteristics of included studies

The eligible articles for this review were published between 2007 and 2024. Only one study was published between 2007 and 2012. However, from 2013 to 2024, there was an increase in number of publications across all continents (Figure 2). This indicated a rising interest in applying genetic methods to distinguish between indigenous and imported cases, which is particularly important for countries approaching zero malaria cases (Dalmat et al. 2019). Africa, Asia, and South America each led with eight publications, followed by North America and Oceania (Table 1). Central America had the fewest, with only one publication. The sample size varied between 45 (Li et al. 2023b) to 8654 subjects (Phelan et al. 2023). There were 15 studies focused on *P. vivax* and 14 on *P. falciparum*. Additionally, four studies investigated both *P. falciparum* and *P. vivax* concurrently. Out of 33 studies, the majority (n=16) focused on a national scale, using genetic data to finely map imported malaria cases within their respective countries. Eight studies conducted global-level analyses, gathering data from around the world. Only five studies were cross-border investigations. Most studies (n=18, 54%) used microsatellite markers. Nine studies used SNP markers/barcodes, and only one combined the data of both microsatellite and SNP markers (Khaireh et al. 2013b). Three studies employed whole genome sequencing (WGS): two for sequencing the entire Plasmodium mitochondrial genome (Schmedes et al. 2019; Li et al. 2023a) and another for sequencing the genome of *P. vivax* (Buyon et al. 2020). We identified four studies employed different approaches for genotyping: one used Single Tandem Repeat Genotyping (Hamedi et al. 2016), while two studies applied Targeted Genome Sequencing of the rif gene (Xu et al. 2023) and the Plasmodium vivax apical membrane antigen-1 (PvAMA-1) gene (Cui et al. 2022). Additionally, one study applied deep sequencing to analyze 35 loci of the *P. falciparum* genome (Holzschuh et al. 2024). Twelve studies included the drug resistance markers in their analysis. Almost half of the studies (n=18, 54%) collected travel survey data using a self-reported questionnaire. Only two studies collected mobile phone data (Tessema et al. 2019). The remaining studies did not explicitly mention collecting travel survey data, mobile data, or GPS tracking data. Regarding the statistical method, the majority of studies used four approaches: (1) estimating expected heterozygosity (HE); (2) calculating Fixation Index (FST) index to investigate the genetic differentiation between populations; (3) Bayesian inference cluster analysis to assess the parasite population structure, and (4) conducting Principal Component Analysis (PCA) to identify population structure and substructure. Detailed characteristics of the included studies are presented in the supplementary material (Table 2).

**Tabel 1.**
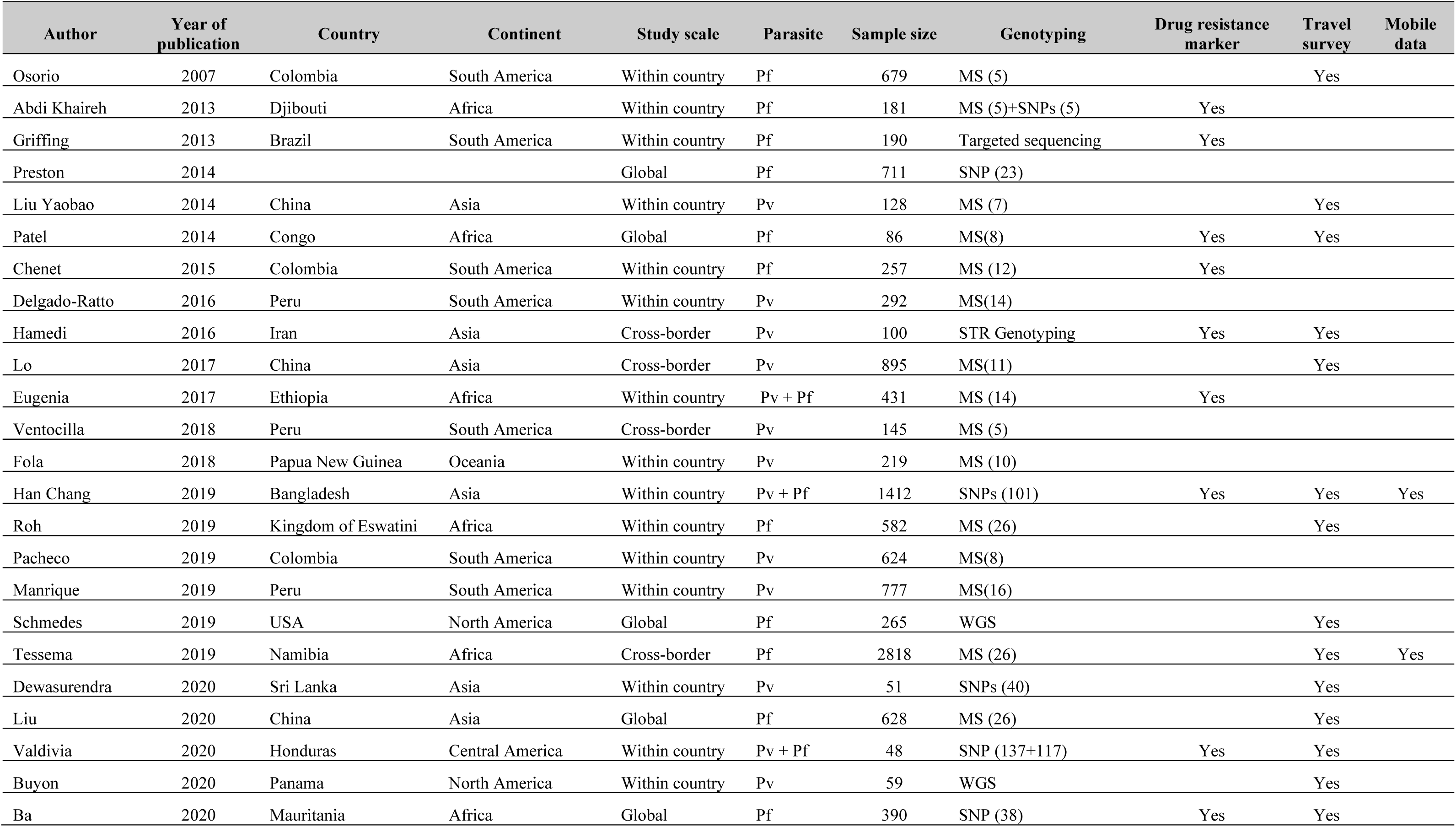
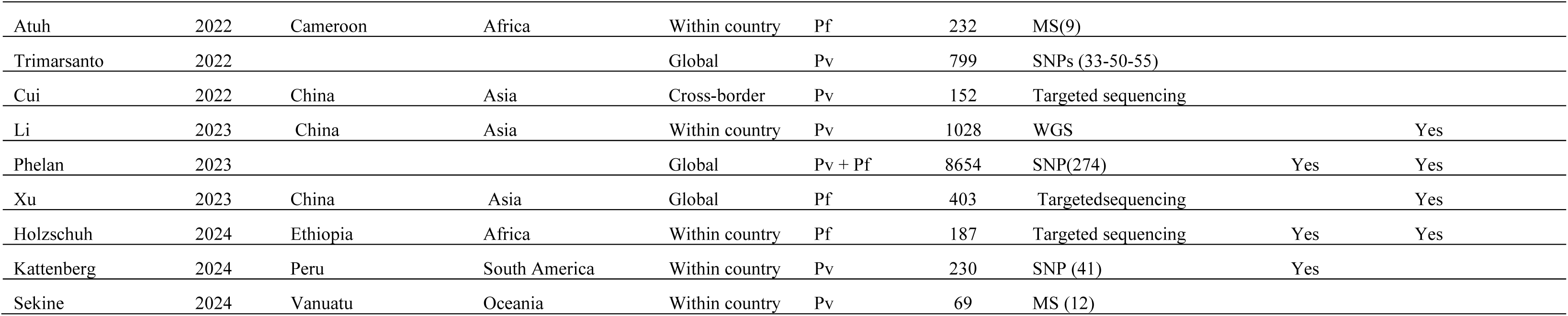
Overview of the eligible studies selected for this review article.

**Figure 2.**
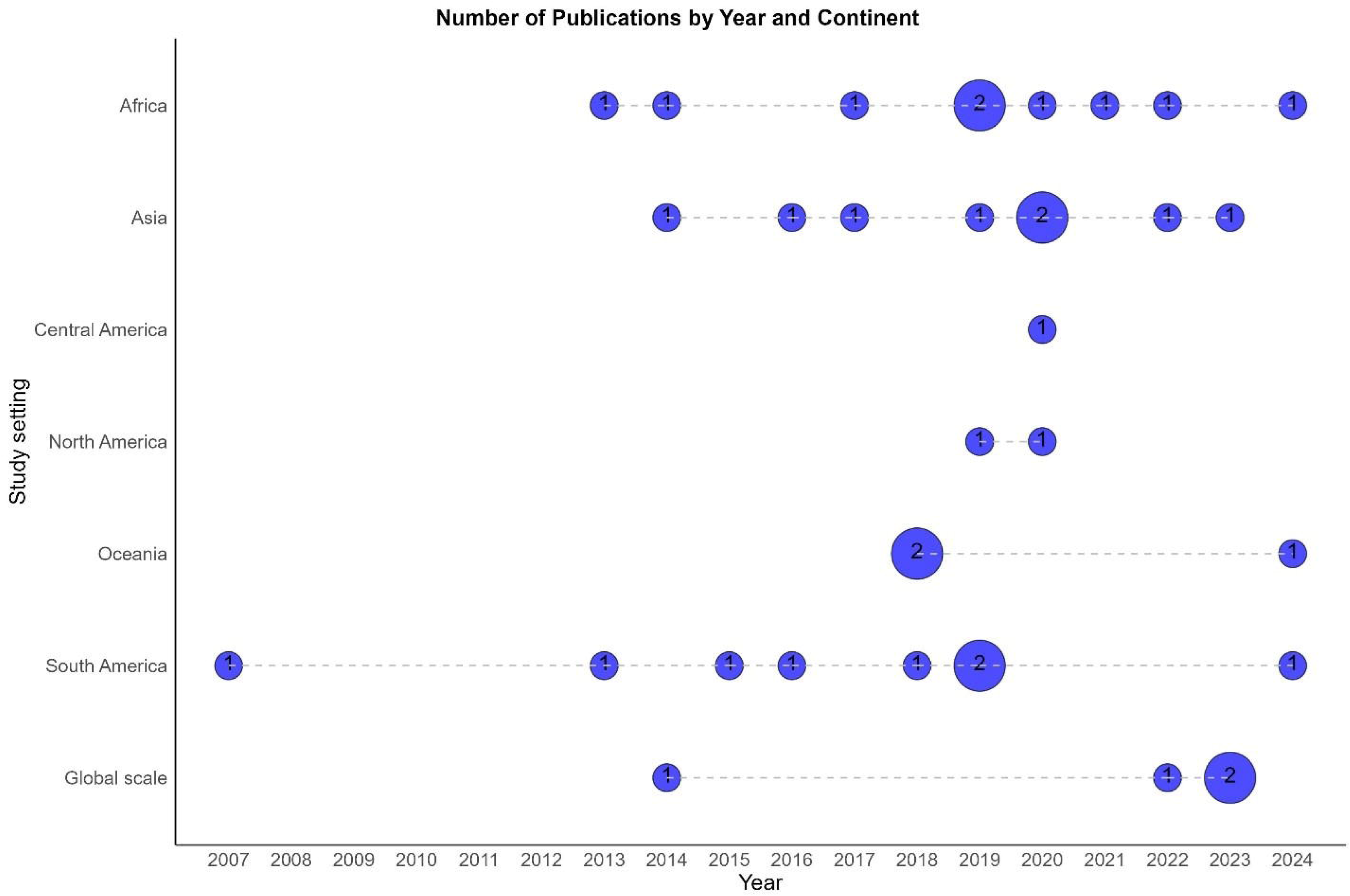
PRISMA flow diagram showing the steps followed to select eligible articles, along with the number of articles retained at each stage

**Tabel 2.**
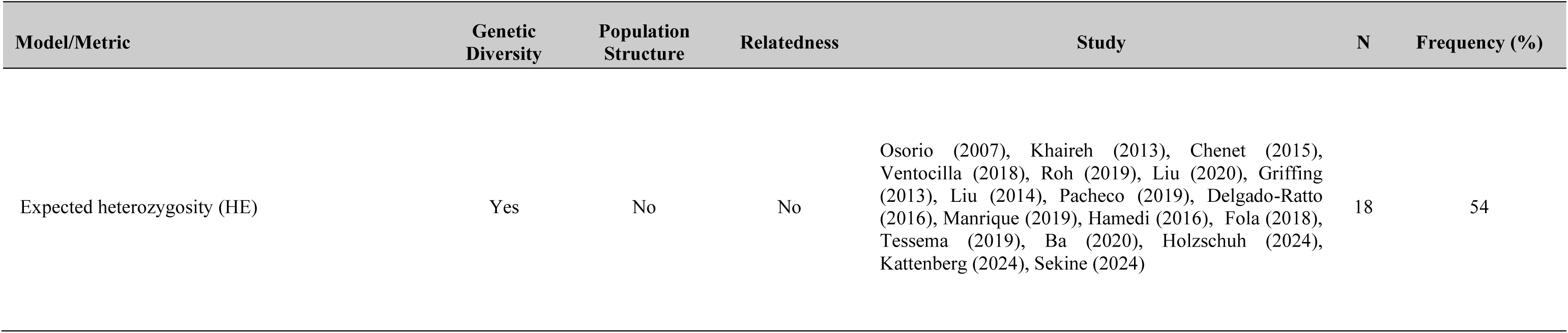
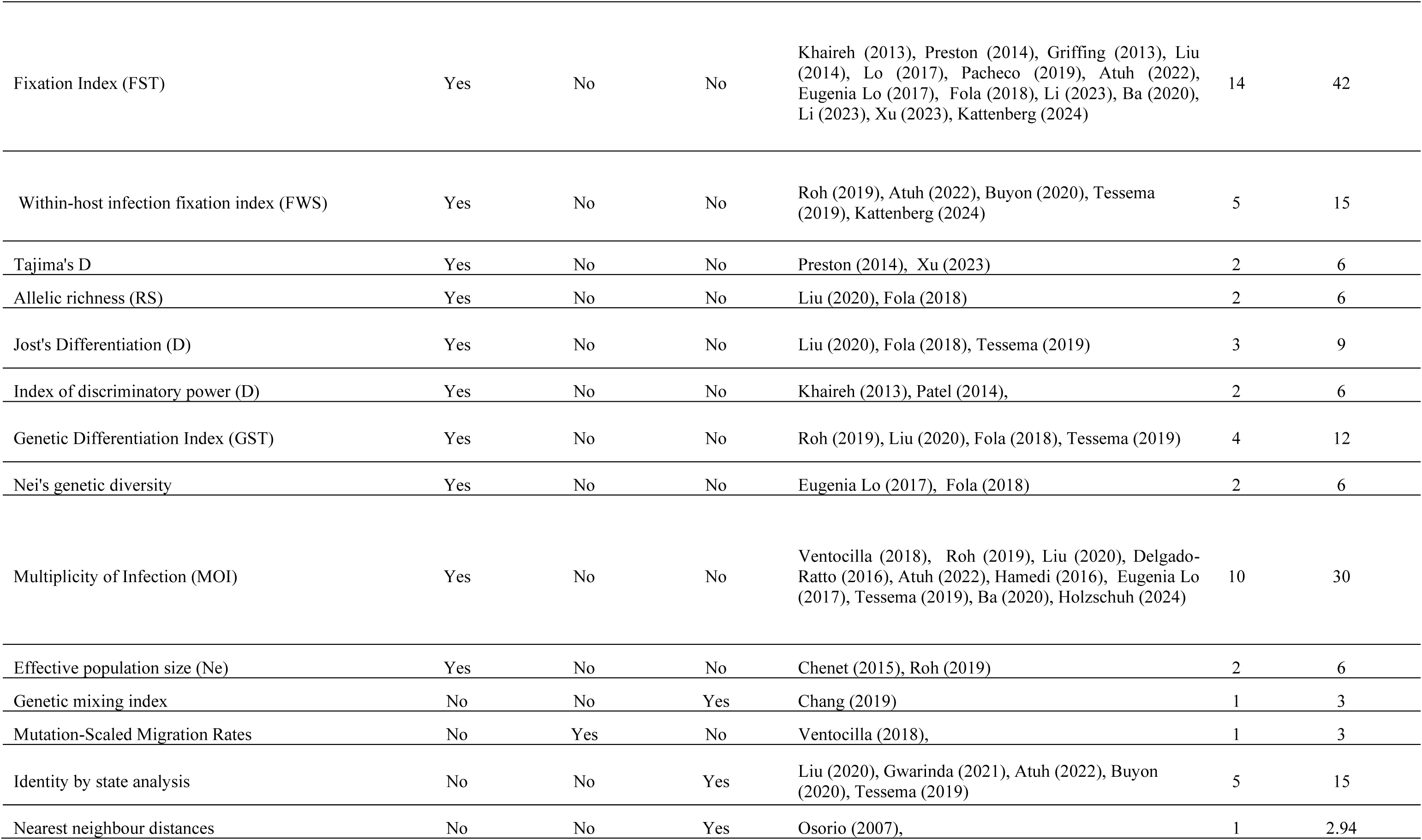
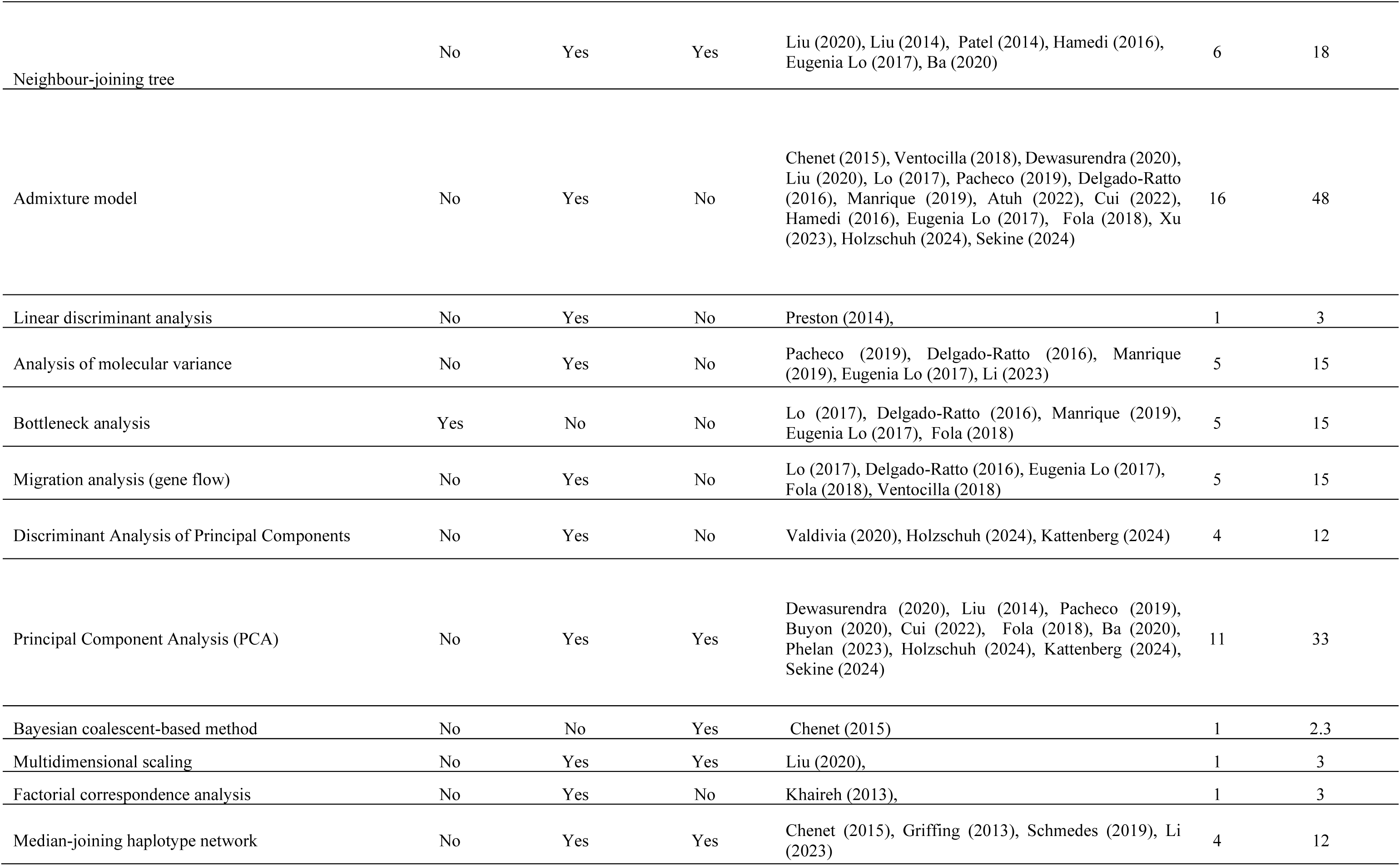
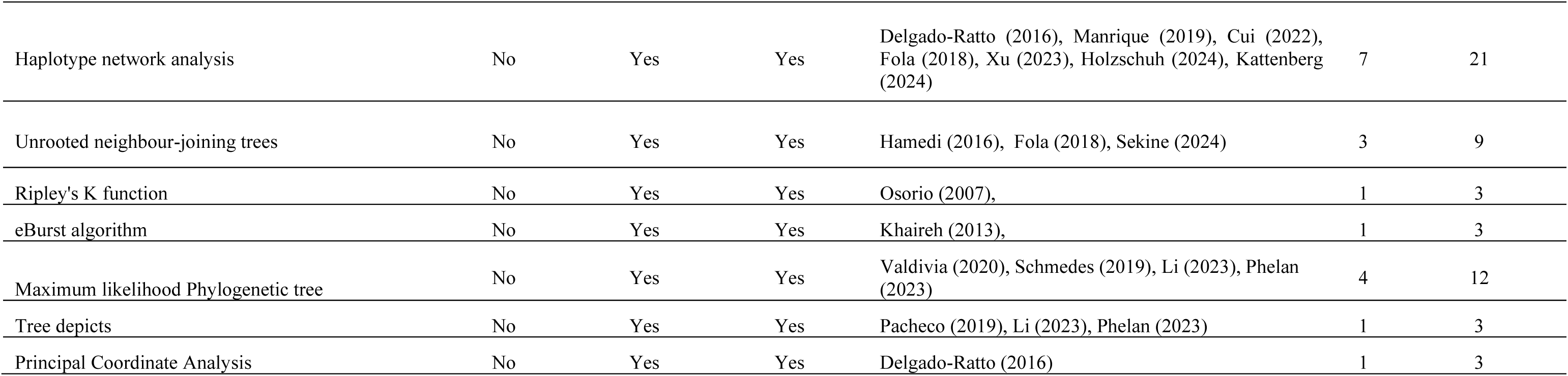
Overview of the analytical methods employed in eligible studies.

### 3.2 Microsatellite markers

Sixteen studies worldwide have investigated the efficiency of MS to determine the genetic diversity and transmission dynamics of malaria parasites. Among these studies, six were conducted in South America: three in Peru and three in Cambodia. The primary focus of studies conducted in Peru was the Amazon region. In 2016, Ventocilla *et al*. analyzed 120 *P. vivax* isolates from the Peruvian North Coast (PNC) using five microsatellite markers. They found low genetic diversity in the PNC but high diversity in the neighboring Ecuadorian Amazon Basin, indicating limited population flow between these regions separated by the Andes Mountains (Ventocilla et al. 2018). In the same year, Delgado-Ratto *et al*. genotyped 292 *P. vivax* isolates from Iquitos and 25 peri-urban and rural villages with 14 microsatellite markers. They identified Iquitos as a reservoir of parasite, spreading genetic variability to the surrounding study areas (Delgado-Ratto et al. 2016). Lastly, Manrique *et al*. conducted molecular surveillance in the Peruvian Amazon using 16 microsatellite markers on 777 mono-infection isolates. Their findings indicated gene flow of *P. vivax*, supporting the hypothesis that human mobility can potentially connect geographically distant areas (Manrique et al. 2019).

Among the studies conducted in Colombia, the earliest was by Osorio *et al*. in 2007. They used five microsatellite to classify 679 *P. falciparum* isolates into imported and indigenous. They found that microsatellite markers had low power to distinguish between these types due to a predominance of a single genotype and low genetic diversity in both imported and indigenous cases (Osorio et al. 2007). Building on this, Chenet *et al*. investigated the genetic composition of 257 *P. falciparum* isolates collected between 2002 and 2009. The findings suggested that fluctuations in the number of cases during this period were likely driven due to local rather than imported cases (Chenet et al. 2015). Expanding the scope to inter-regional human migration, Pacheco *et al*. (2019) evaluated the genetic differentiation of 624 *P. vivax* isolates across four geographically separated areas using eight microsatellite markers. Contrary to expectations of clear geographic structure, the study revealed only moderate genetic differentiation. This suggested a corridor between the northwest and the south Pacific Coast of Colombia, where migration of infected individuals enabled the spread of the parasite (Pacheco et al. 2018).

The primary focus of studies published in Africa was on Central Africa. Patel *et al*. (2014) investigated whether microsatellite markers could identify the origin of malaria outbreaks among United Nations soldiers returning from the Democratic Republic of the Congo (DRC) to Guatemala. Using eight microsatellite markers, they found that the parasites in the soldiers were genetically similar to those from the DRC, suggesting the outbreak was likely due to *P. falciparum* imported cases from the DRC (Patel et al. 2014). In 2019, Roh *et al*. focused on distinguishing between imported and locally acquired *P. falciparum* infections in Eswatini. They genotyped 582 isolates using 26 microsatellite markers and found nearly identical levels of genetic diversity in both imported and local infections, making it challenging to differentiate between the two types of infections (Roh et al. 2019). In comparison, Tessema *et al*. integrated travel history, mobile phone data, and parasite genetics to study malaria connectivity within Namibia and across the Angolan and Zambian borders. By genotyping 4,643 *P. falciparum* isolates using 26 markers, they provided strong evidence that the majority of cases in northeastern Namibia were attributed to local transmission. This finding was consistent with estimates derived from mobile phone and travel history data (Tessema et al. 2019). Atuh *et al*. (2022) examined the genetic connectivity of *P. falciparum* in Northwestern and Southwestern Cameroon using nine microsatellite markers. Despite high human migration between the regions, their analysis of 232 isolates revealed only a small degree of genetic similarity, indicating limited gene flow between these areas (Atuh et al. 2022). In contrast to previous studies, Eugenia Lo *et al*. used 14 microsatellites to assess gene flow patterns of both *P. falciparum* (N = 226) and *P. vivax* (N = 205) isolates in Ethiopia. The findings revealed that human migrations may promote parasites gene flow between the northern and eastern regions of the country(Lo, Hemming-Schroeder, et al. 2017).

We identified three studies conducted in Asia, all carried out in China. Yaobao Liu *et al*. (2014) analyzed 128 *P. vivax* isolates from Central China using seven microsatellite markers. Despite low endemicity, they found high genetic diversity, indicating gene flow among central provinces. However, the markers were not effective in distinguishing local from imported infections, leaving the sources of this diversity unclear (Liu et al. 2014). Similarly, Lo *et al*. (2017) examined the genetic diversity of *P. vivax* across 895 samples from the Myanmar-China border using 11 microsatellite markers. They also observed high genetic diversity, suggesting that human migration facilitated parasite gene flow both locally and across the border (Lo, Lam, et al. 2017). In contrast, Liu *et al*. (2020) focused on identifying the geographic origin of imported *P. vivax* infections. They analyzed 602 cases traveled from 26 sub-Saharan African countries to China using 26 microsatellite markers. Genetically related infections were found in people who traveled to the same country at about the same time. The findings demonstrated the potential of these markers to trace the origin of infections on a global scale (Liu et al. 2020b).

Only two studies were conducted in Oceania, reflecting the limited research carried out on this continent compared to others. To understanding the migration patterns of *P. vivax* in Papua New Guinea (PNG), Fola *et al*. genotyped 219 *P. vivax* isolates suing ten microsatellite markers. The study identified three major genetic populations, with gene flow patterns aligning with human migration routes. Notably, gene flow was higher among Mainland parasite than among Island populations (Fola et al. 2018). In 2024, Sekine *et al*. Traced the origins of *P. vivax* among 69 cases from Aneityum Island in Vanuatu. The results of genotyping with five microsatellite markers showed that the parasites responsible for the outbreak in 2002 were imported. However, the exact source(s) could not be determined (Sekine et al. 2024).

### 3.3 SNP markers (SNPs)

We found eight studies that used SNP markers to assess the geographic origin of malaria infections, with two of these studies conducted in Asia. In Sri Lanka, Dewasurendra *et al*. (2016) investigated *P. vivax* origins using a 40-SNP barcode on samples collected from 2005 to 2011. Nine samples clustered with the South American group, indicating these isolates were most probably imported cases. However, the lack of travel histories made such inferences remain unconfirmed (Dewasurendra et al. 2020). In contrast, Chang *et al*. (2019) integrated data from a 101-SNP barcode with the travel survey and mobile phone data to assess malaria transmission countrywide in Bangladesh. They could detect likely imported cases among 1412 malaria-positive cases (Chang, Wesolowski, Sinha, Jacob, Mahmud, Uddin, Zaman, Hossain, Faiz, Ghose, et al. 2019).

We identified a study conducted in Peru, South America by Kattenberg *et al*. They developed a 41-SNP barcode, specifically for *P. vivax* samples from the Peruvian Amazon. Seven out of 230 samples (3%) were predicted to originate from Vietnam, Afghanistan, and Iran. However, their findings could not be fully confirmed as incorrect predictions might have arisen due to missing data in SNP barcode (Kattenberg et al. 2024). The same approach was applied by Ba *et al*. to compare *P. vivax* population in Mauritania, West Africa, with parasite populations in other countries. The results of a 38-SNP barcode revealed that the Mauritanian samples (n=100) formed a distinct geographical group, with slightly closer relatedness to Ethiopia than to the samples from Southeast Asia (Ba et al. 2020). In Central America, only one study in Honduras explored the population structure of both *P. vivax* and *P. falciparum* isolates. The results showed that two out of three isolates with chloroquine-resistant parasites were imported cases from Africa. In contrast, SNP barcodes could not provide any information about the origin of infection for *P. vivax* (Valdivia et al., 2020).

Lastly, we found three studies conducted on a global scale. The first study, published by Preston *et al*. in 2014, included 711 *P. falciparum* isolates from 14 countries and developed a 23-SNP barcode. The SNP barcode determined the geographic origin of parasite isolates with 92% accuracy, as confirmed by available data regarding the origin of each sample (Preston et al. 2014). In comparison, Trimarsanto *et al*. (2022) used a larger number of SNPs (n=33) to predict the infection origin of 799 *P. vivax* isolates collected from 21 countries. The findings confirmed the capacity of this SNP panel to predict infection’s country of origin accurately (Trimarsanto et al. 2022). The most recent publication was conducted by Phelan *et al*. in 2023. They used the largest number of genetic markers, incorporating a combination of drug resistance mutations, mitochondrial SNPs, and a set of established SNP markers. Analyzing data from 7,152 *P. falciparum* and 1,502 *P. vivax* samples revealed that genetic markers could predict the geographical origin of infections, with accuracy rates of 96.1% at the continental level and 94.6% at the regional level (Phelan et al. 2023).

### 3.4 SNPs and Microsatellite markers

Khaireh *et al*. genotyped five microsatellites and five SNPs associated with pyrimethamine resistance in 185 *P. falciparum* isolates collected over an 11-year period (1998, 1999, 2002, and 2009) from the low malaria transmission setting of Djibouti. The most notable finding was the decline in genetic diversity among parasite populations over the study period. This might be attributed to a decline in malaria transmission from neighboring countries, particularly Ethiopia (Khaireh et al. 2013a).

### 3.5 Targeted sequencing

We identified five studies focused on targeted genome sequencing. Cui *et al*. investigated the genetic diversity of the P. vivax apical membrane antigen-1 (PvAMA-1) gene, an important vaccine candidate for vivax malaria, along the China–Myanmar border, including 152 imported cases in China and 73 isolates from Myanmar. They found high genetic diversity and positive selection in the PvAMA-1 gene among imported cases, implying that traveling may affect the population structure and genetic characteristics of malaria on the Chinese side of the China–Myanmar border (Cui et al. 2022). Xu *et al*. (2023) studied nucleotide diversity in the polymorphic region of the rif gene which is involved in immune evasion. High genetic differentiation was found between 53 malaria cases imported from Ghana to China and South Asian populations, including, Thailand, Vietnam, Myanmar, and Cambodia. The findings indicated that the P*. falciparum* populations have a substantial continental genetic structure (Xu et al. 2023). Unlike other studies, Hamedi *et al*. genotyped nine Single Tandem Repeat (STR) markers on the pvmdr1 gene, which is associated with drug resistance, particularly in *P. vivax*. They could not find any notable genetic differentiation between 23 imported cases from Afghanistan or Pakistan and 73 autochthonous cases in Iran (Hamedi et al. 2016). In Brazil, Griffing *et al*. genotyped blood spots collected on filter paper from the 1980s and 1990s across three regions to investigate internal migration of *P. falciparum*. They analyzed 190 samples using 28 microsatellite markers on the dhfr and dhps genes, known to be associated with sulphadoxine/pyrimethamine (SP) resistance. Their findings suggested that internal migration of the parasite between the regions might lead to admixture of parasite lineages (Griffing et al. 2013). In one of the most recent studies, Holzschuh *et al*. sequenced 187 *P. falciparum* samples from two regions in Ethiopia. Unlike other research, they used multiplex amplicon deep sequencing to analyze 35 loci, including those associated with drug resistance. The findings suggested that individuals with recent travel history might initiate local transmission of the malaria parasite within the community which then led to ongoing local transmission (Holzschuh et al. 2024).

### 3.6 Whole genome sequencing data

Only three studies used whole genome sequencing data with different approaches: two focused on the mitochondrial genome of *P. falciparum* (Schmedes et al. 2019) and *P. vivax* (Li et al. 2023a), while the other examined the whole genome of *P. vivax* (Buyon et al. 2020). In 2019, Schmedes *et al*. conducted sequencing of the Plasmodium mitochondrial genome to determine the geographical origin of imported *P. falciparum* parasites to the United States. The study included 265 imported malaria cases between 2014 and 2017. The genetic data could successfully determine the origin of 13 samples in specific regions or countries like the Philippines and Ghana. However, for most samples, geographical origin prediction was not possible due to common mitochondrial haplotypes present worldwide (Schmedes et al. 2019). The same approach was applied by Li *et al*. to investigate the geographical origin of *P. vivax* cases in Hainan, China. The origin of seven out of 14 imported cases could be inferred which was consistent with recorded travel histories. However, the origin of other imported cases could not be traced, raising questions about the suitability of using mitochondrial genomes in this context (Li et al. 2023b). In 2020, Buyon *et al*. explored patterns of recent common ancestry among 59 *P. vivax* samples from Panama collected from 2007–2009 and 2017–2019. The results showed that four samples with travel history did not exhibit recent ancestry connections with other Panamanian samples; instead, they formed a cluster with samples collected in a previous study from Colombia. Nevertheless, there was no indication of outcrossing between the potentially imported parasites and the local Panamanian parasite population (Buyon et al. 2020).

### 3.7 Statistical models

The methods can be broadly categorized into three main groups: those that assess genetic diversity, population structure, and relatedness among populations or individuals. The most common methods were determining genetic diversity and population structure. Regarding the assessment of genetic diversity, expected heterozygosity (HE) was the most used method applied in 18 studies, accounting for 54% of the total. This metric was primarily used to indirectly measure the impact of imported cases on genetic variation within the local populations, specifically in the context of longitudinal studies. Such studies measured HE over a defined period, allowing them to identify and analyze trends in HE metric changes over time. The Fixation Index (FST) was the second most frequently used method, appearing in 14 studies (42%). Like HE, FST was used to measure the genetic differentiation among populations, in some cases, extrapolating the FST as measure of gene flow. The Admixture model was used in 16 studies (48%) to analyze population structure, providing insights into the extent of genetic mixing resulting from migration between different populations. Principal Component Analysis (PCA) was employed in 11 studies (33 %). PCA was used to detect population structure and substructure (Table 2).

Interestingly, we found only five studies (15%) that applied methods capable of directly estimating potential evidence of migration between populations. Delgado-Ratto *et al*. (2016) assessed human migration patterns in and around Iquitos City using a Bayes approach based on the coalescence theory. The findings showed that such a model can estimate parasite importation’s magnitude and direction. Unfortunately, no information on travel patterns was collected in that study (Delgado-Ratto et al. 2016). To explore gene flow between the Peruvian North Coast and the Ecuadorian Amazon Basin, Ventocilla *et al*. developed two migration models using a Bayesian approach with MIGRATE-N. The models showed the presence of three distinct subpopulations with some level of migration occurring between them. These findings were consistent with the results obtained from the structure analysis (Ventocilla et al. 2018). In a similar approach, Eugenia Lo *et al*. estimated the intensity and direction of gene flow using a Bayesian approach implemented in BayesAss program. The bidirectional migration was observed between the north and east Ethiopia, indicating that human migrations may facilitate the parasite gene flow (Lo, Hemming-Schroeder, et al. 2017). In 2017, Lo *et al*. applied a comparable method, using a Bayesian approach to estimate the posterior probability distribution of the proportion of migrants from one population to another. The findings revealed a greater migration rate from Myanmar to China. By contrast, migration from China to Myanmar was relatively small (Lo, Lam, et al. 2017). Unlike the other studies, Fola *et al*. (2018) used divMigrate-online software to estimate migration patterns among different geographic regions of Papua New Guinea. Their findings showed more migration within the mainland than within the Island region and relatively limited migration between them (Fola et al. 2018).

In addition, we identified two studies that developed a novel method to determine if an isolate is imported. Chang *et al*. compared the geographic distance between isolates with geographic distances estimated based on genetic data. They quantified the probability of a geographic distance given an SNP difference by applying Bayes’ rule (Chang, Wesolowski, Sinha, Jacob, Mahmud, Uddin, Zaman, Hossain, Faiz, Ghose, et al. 2019). In contrast, Tessema *et al*. developed an approach that combined parasite genetic data and travel history data to estimate local and cross-border importation in Namibia. Infections from Namibia were more found to be genetically related to those from southern Angola and Zambia, compared to northern Angola. This indicated parasite mixing within the geographically connected Namibia-Angola-Zambia regional zone (Tessema et al. 2019).

## 4 Discussion

Understanding the spatial spread of malaria parasites, particularly in low transmission settings where imported cases can undermine local interventions, is crucial for effective control and elimination efforts (Dalmat et al. 2019). This review synthesizes evidence on the routine use of parasite genetic data to map imported malaria, offering insights into methods and approaches for national control programs and elimination strategies.

This review highlights the diverse methods used to analyze parasite genetic data for mapping imported malaria cases, emphasizing the strengths and weaknesses of each approach. The findings indicated that molecular techniques such as microsatellite genotyping can offer valuable insights into parasite population structure and migration patterns. These markers have been widely used to study genetic diversity and population dynamics in various settings, including the Peruvian Amazon (Delgado-Ratto et al. 2016), Central Africa (Patel et al. 2014), Asia (Liu et al. 2020a), and Papua New Guinea (Fola et al. 2018). The findings revealed patterns of gene flow and population differentiation, highlighting the role of human migration in shaping malaria transmission. However, these studies suffered from limited discriminatory power in regions with low genetic diversity and challenges distinguishing imported cases from local transmission in areas with closely related parasite clones (Roh et al. 2019).

Similarly, SNP markers have been employed to identify the geographic origin of infections. Studies in diverse settings such as Sri Lanka, Bangladesh, and Honduras have used SNP barcodes to track parasite movement and assess population structure. The findings of such studies demonstrated the accuracy of SNP markers in classifying imported infections at both regional and global levels (Phelan et al. 2023). It is worth mentioning that, like microsatellite markers, SNP markers also lack sufficient resolution to discriminate between imported and local parasites on a smaller geographical scale, such as within a country, limiting their application (Valdivia et al. 2020).

Compared to microsatellite and SNP barcodes, whole genome sequencing (WGS) offers a comprehensive approach to understanding the genetic makeup of malaria parasites, providing insights into their origin, transmission dynamics, and evolution. Its ability to analyze the entire genome allows for a more detailed assessment of parasite diversity and population structure than other molecular techniques (Akoniyon et al. 2022). WGS provides high-resolution data, enabling researchers to identify genetic variations at the nucleotide level, including single nucleotide polymorphisms (SNPs), insertions, and deletions. This level of detail is invaluable for tracking the spread of parasite populations, identifying transmission routes, and detecting emerging drug resistance (Winter et al. 2015). Moreover, WGS can reveal fine-scale population structure, helping to differentiate between closely related parasite strains and accurately trace their origins. Additionally, WGS data can be combined with epidemiological and clinical information to provide a comprehensive understanding of malaria transmission dynamics, identify the primary sources of malaria importation, and assess the impact of this importation on the genetic structure of local parasite populations. Such insights can subsequently be used to inform and optimize targeted intervention strategies (Neafsey et al. 2021).

While WGS is considered a promising approach, it presents particular challenges in accurately determining the geographic origin of imported parasites. One fundamental limitation is the presence of common haplotypes, which can reduce the predictive accuracy of WGS analysis. Additionally, the infrastructure required for WGS analysis, including specialized equipment and expertise, presents a significant barrier to widespread implementation. Moreover, the high cost associated with WGS makes it impractical for routine use in all regions affected by malaria. However, addressing these challenges will require concerted efforts and investment in infrastructure and research capabilities. It is important to acknowledge that, despite its many advantages, whole genome sequencing (WGS) should not be relied upon exclusively. A well-designed study framework that considers the specific study settings and populations is essential. This framework should ensure that samples are collected, transported, and analyzed efficiently to maintain their integrity. Additionally, it must integrate genetic data with information from traveler surveys and epidemiological studies, allowing for a comprehensive understanding of malaria transmission dynamics and the impact of malaria importation on local populations.

The findings revealed that current approaches for determining imported malaria cases have limitations in accurately tracing changes in parasite genetics over time. These methods often rely on cross-sectional data, which provide snapshots of parasite diversity at specific time points but may not capture the full extent of parasite transmission dynamics (Collins and Duffy 2022). To address this limitation, future research should focus on longitudinal studies that track parasite movement over time. Researchers can better understand how parasites spread and evolve in response to changing environmental conditions and control interventions by analyzing genetic data from sequential time points. In addition, longitudinal analysis will enable researchers to assess the effectiveness of malaria control measures and identify transmission hotspots with potential imported parasites (Kwiatkowski 2024). Ultimately, longitudinal studies will be essential for developing targeted interventions and evaluating their impact on malaria transmission dynamics. A comprehensive understanding of the study population’s behavior and setting characteristics is necessary to plan the research activities.

Utilizing state-of-the-art sequencing technology alongside advanced statistical techniques, such as machine learning methods, can hold promise for enhancing the analysis of parasite genetic data and deepening our understanding of malaria transmission dynamics. Machine learning algorithms can efficiently process large-scale genomic datasets to identify complex patterns associated with parasite migration and population structure (Golumbeanu et al. 2022). However, integrating machine learning methods into malaria research requires careful consideration of data quality, model interpretation, and computational resources. Future studies should focus on refining machine learning algorithms and validating their capabilities in diverse malaria-endemic settings to maximize their utility in informing evidence-based control and elimination strategies (Mbunge et al. 2023).

## Supporting information

Appendix 1: Search commands used in various databases to identify relevant articles

## 7 Author Contributions

Conceptualization: MS, CDR. Data curation: MS, LCS. Formal analysis: MS. Funding acquisition: CDR, DG, JPV. Investigation: MS, LCS, CDR. Resources: MS, CDR. Supervision: CDR. Validation: LCS, CDR. Visualization: MS. Writing – original draft: MS. Writing – review & editing: LCS, DG, CDR, JPV. All authors approved the final version of the manuscript

## 9 Acknowledgments

NA

## 10 Data Availability Statement

NA

## References

Akoniyon, Olusegun Philip, Taiye Samson Adewumi, Leah Maharaj, Olukunle Olugbenle Oyegoke, Alexandra Roux, Matthew A Adeleke, Rajendra Maharaj, and Moses Okpeku. 2022. “Whole genome sequencing contributions and challenges in disease reduction focused on malaria.” Biology 11 (4):587.

Ariey, Frédéric, Jean-Bernard Duchemin, and Vincent Robert. 2003. “Metapopulation concepts applied to falciparum malaria and their impacts on the emergence and spread of chloroquine resistance.” Infection, Genetics and Evolution 2 (3):185–192.

Atuh, N. I., D. N. Anong, F. C. Jerome, E. Oriero, N. I. Mohammed, U. D’Alessandro, and A. Amambua-Ngwa. 2022. “High genetic complexity but low relatedness in Plasmodium falciparum infections from Western Savannah Highlands and coastal equatorial Lowlands of Cameroon.” Pathogens and Global Health 116 (7):428–437. doi: 10.1080/20477724.2021.1953686.

Ba, H., S. Auburn, C. G. Jacob, S. Goncalves, C. W. Duffy, L. B. Stewart, R. N. Price, Y. B. Deh, M. Y. Diallo, A. Tandia, D. P. Kwiatkowski, and D. J. Conway. 2020. “Multi-locus genotyping reveals established endemicity of a geographically distinct Plasmodium vivax population in Mauritania, West Africa.” PLoS Negl Trop Dis 14 (12):e0008945. doi: 10.1371/journal.pntd.0008945.

Buckee, Caroline O, Amy Wesolowski, Nathan N Eagle, Elsa Hansen, and Robert W Snow. 2013. “Mobile phones and malaria: modeling human and parasite travel.” Travel medicine and infectious disease 11 (1):15–22.

Buyon, L. E., A. M. Santamaria, A. M. Early, M. Quijada, I. Barahona, J. Lasso, M. Avila, S. K. Volkman, M. Marti, D. E. Neafsey, and N. Obaldia Iii. 2020. “Population genomics of Plasmodium vivax in Panama to assess the risk of case importation on malaria elimination.” PLoS Negl Trop Dis 14 (12):e0008962. doi: 10.1371/journal.pntd.0008962.

Carrasco-Escobar, Gabriel, Kimberly Fornace, Daniel Wong, Pierre G Padilla-Huamantinco, Jose A Saldaña-Lopez, Ober E Castillo-Meza, Armando E Caballero-Andrade, Edgar Manrique, Jorge Ruiz-Cabrejos, and Jose Luis Barboza. 2020. “Open-source 3D printable GPS tracker to characterize the role of human population movement on malaria epidemiology in river networks: a proof-of-concept study in the Peruvian Amazon.” Frontiers in Public Health 8:526468.

Chang, H. H., A. Wesolowski, I. Sinha, C. G. Jacob, A. Mahmud, D. Uddin, S. I. Zaman, M. A. Hossain, M. A. Faiz, A. Ghose, A. A. Sayeed, M. R. Rahman, A. Islam, M. J. Karim, M. K. Rezwan, A. K. M. Shamsuzzaman, S. T. Jhora, M. M. Aktaruzzaman, E. Drury, S. Gonçalves, M. Kekre, M. Dhorda, R. Vongpromek, O. Miotto, K. Engø-Monsen, D. Kwiatkowski, R. J. Maude, and C. Buckee. 2019. “Mapping imported malaria in Bangladesh using parasite genetic and human mobility data.” eLife 8. doi: 10.7554/eLife.43481.

Chang, HH, A Wesolowski, I Sinha, CG Jacob, A Mahmud, D Uddin, SI Zaman, MA Hossain, MA Faiz, and A Ghose. 2019. Mapping imported malaria in Bangladesh using parasite genetic and human mobility data. Elife 8.

Chenet, S. M., J. E. Taylor, S. Blair, L. Zuluaga, and A. A. Escalante. 2015. “Longitudinal analysis of Plasmodium falciparum genetic variation in Turbo, Colombia: implications for malaria control and elimination.” Malar J 14:363. doi: 10.1186/s12936-015-0887-9.

Collins, OC, and KJ Duffy. 2022. “A mathematical model for the dynamics and control of malaria in Nigeria.” Infectious disease modelling 7 (4):728–741.

Cui, Y. B., H. M. Shen, S. B. Chen, K. Kassegne, T. Q. Shi, B. Xu, J. H. Chen, J. H. Wu, and Y. Wang. 2022. “Genetic Diversity and Selection of Plasmodium vivax Apical Membrane Antigen-1 in China–Myanmar Border of Yunnan Province, China, 2009–2016.” 11. doi: 10.3389/fcimb.2021.742189.

Dalmat, Ronit, Brienna Naughton, Tao Sheng Kwan-Gett, Jennifer Slyker, and Erin M Stuckey. 2019. “Use cases for genetic epidemiology in malaria elimination.” Malaria journal 18:1–11.

Delgado-Ratto, Christopher, Dionicia Gamboa, Veronica E Soto-Calle, Peter Van den Eede, Eliana Torres, Luis Sanchez-Martinez, Juan Contreras-Mancilla, Anna Rosanas-Urgell, Hugo Rodriguez Ferrucci, and Alejandro Llanos-Cuentas. 2016. “Population genetics of Plasmodium vivax in the Peruvian Amazon.” PLoS neglected tropical diseases 10 (1):e0004376.

Dewasurendra, R. L., M. L. Baniecki, S. Schaffner, Y. Siriwardena, J. Moon, R. Doshi, S. Gunawardena, R. F. Daniels, D. Neafsey, S. Volkman, N. V. Chandrasekharan, D. F. Wirth, and N. D. Karunaweera. 2020. “Use of a Plasmodium vivax genetic barcode for genomic surveillance and parasite tracking in Sri Lanka.” Malar J 19 (1):342. doi: 10.1186/s12936-020-03386-3.

Feachem, Richard GA, Ingrid Chen, Omar Akbari, Amelia Bertozzi-Villa, Samir Bhatt, Fred Binka, Maciej F Boni, Caroline Buckee, Joseph Dieleman, and Arjen Dondorp. 2019. “Malaria eradication within a generation: ambitious, achievable, and necessary.” The Lancet 394 (10203):1056–1112.

Fola, A. A., E. Nate, G. L. Abby Harrison, C. Barnadas, M. W. Hetzel, J. Iga, P. Siba, I. Mueller, and A. E. Barry. 2018. “Nationwide genetic surveillance of Plasmodium vivax in Papua New Guinea reveals heterogeneous transmission dynamics and routes of migration amongst subdivided populations.” Infect Genet Evol 58:83–95. doi: 10.1016/j.meegid.2017.11.028.

Golumbeanu, Monica, Guo-Jing Yang, Flavia Camponovo, Erin M Stuckey, Nicholas Hamon, Mathias Mondy, Sarah Rees, Nakul Chitnis, Ewan Cameron, and Melissa A Penny. 2022. “Leveraging mathematical models of disease dynamics and machine learning to improve development of novel malaria interventions.” Infectious Diseases of Poverty 11 (03):37–53.

Griffing, S. M., G. M. R. Viana, T. Mixson-Hayden, S. Sridaran, M. T. Alam, A. M. de Oliveira, J. W. Barnwell, A. A. Escalante, M. M. Povoa, and V. Udhayakumar. 2013. “Historical Shifts in Brazilian P. falciparum Population Structure and Drug Resistance Alleles.” PLoS ONE 8 (3). doi: 10.1371/journal.pone.0058984.

Hamedi, Y., K. Sharifi-Sarasiabi, F. Dehghan, R. Safari, S. To, I. Handayuni, H. Trimarsanto, R. N. Price, and S. Auburn. 2016. “Molecular Epidemiology of P. vivax in Iran: High Diversity and Complex Sub-Structure Using Neutral Markers, but No Evidence of Y976F Mutation at pvmdr1.” PLoS One 11 (11):e0166124. doi: 10.1371/journal.pone.0166124.

Holzschuh, A., Y. Ewnetu, L. Carlier, A. Lerch, I. Gerlovina, S. C. Baker, D. Yewhalaw, W. Haileselassie, N. Berhane, W. Lemma, and C. Koepfli. 2024. “Plasmodium falciparum transmission in the highlands of Ethiopia is driven by closely related and clonal parasites.” Mol Ecol 33 (6):e17292. doi: 10.1111/mec.17292.

Kattenberg, J. H., L. Cabrera-Sosa, E. Figueroa-Ildefonso, M. Mutsaers, P. Monsieurs, P. Guetens, B. Infante, C. Delgado-Ratto, D. Gamboa, and A. Rosanas-Urgell. 2024. “Plasmodium vivax genomic surveillance in the Peruvian Amazon with Pv AmpliSeq assay.” PLoS Negl Trop Dis 18 (7):e0011879. doi: 10.1371/journal.pntd.0011879.

Khaireh, B. A., A. Assefa, H. H. Guessod, L. K. Basco, M. A. Khaireh, A. Pascual, S. Briolant, S. M. Bouh, I. H. Farah, H. M. Ali, A. I. A. Abdi, M. O. Aden, Z. Abdillahi, S. N. Ayeh, H. Y. Darar, J. L. Koeck, C. Rogier, B. Pradines, and H. Bogreau. 2013a. “Population genetics analysis during the elimination process of Plasmodium falciparum in Djibouti.” Malaria Journal 12 (1). doi: 10.1186/1475-2875-12-201.

Khaireh, B. A., A. Assefa, H. H. Guessod, L. K. Basco, M. A. Khaireh, A. Pascual, S. Briolant, S. M. Bouh, I. H. Farah, H. M. Ali, A. I. Abdi, M. O. Aden, Z. Abdillahi, S. N. Ayeh, H. Y. Darar, J. L. Koeck, C. Rogier, B. Pradines, and H. Bogreau. 2013b. “Population genetics analysis during the elimination process of Plasmodium falciparum in Djibouti.” Malar J 12:201. doi: 10.1186/1475-2875-12-201.

Kwiatkowski, Dominic. 2024. “Modelling transmission dynamics and genomic diversity in a recombining parasite population.” Wellcome Open Research 9:215.

Li, Y., X. Huang, L. Qing, W. Zeng, X. Zeng, F. Meng, G. Wang, and Y. Chen. 2023a. “Geographical origin of Plasmodium vivax in the Hainan Island, China: insights from mitochondrial genome.” Malar J 22 (1):84. doi: 10.1186/s12936-023-04520-7.

Li, Y., X. Huang, L. Qing, W. Zeng, X. Zeng, F. Meng, G. Z. Wang, and Y. Chen. 2023b. “Geographical origin of Plasmodium vivax in the Hainan Island, China: insights from mitochondrial genome.” Malaria Journal 22 (1). doi: 10.1186/s12936-023-04520-7.

Liu, Y., S. Auburn, J. Cao, H. Trimarsanto, H. Zhou, K. A. Gray, T. G. Clark, R. N. Price, Q. Cheng, R. Huang, and Q. Gao. 2014. “Genetic diversity and population structure of Plasmodium vivax in Central China.” Malar J 13:262. doi: 10.1186/1475-2875-13-262.

Liu, Y. B., S. K. Tessema, M. Murphy, S. Xu, A. Schwartz, W. M. Wang, Y. Y. Cao, F. Lu, J. X. Tang, Y. P. Gu, G. D. Zhu, H. Y. Zhou, Q. Gao, R. Huang, J. Cao, and B. Greenhouse. 2020a. “Confirmation of the absence of local transmission and geographic assignment of imported *falciparum* malaria cases to China using microsatellite panel.” Malaria Journal 19 (1):11. doi: 10.1186/s12936-020-03316-3.

Liu, Y., S. K. Tessema, M. Murphy, S. Xu, A. Schwartz, W. Wang, Y. Cao, F. Lu, J. Tang, Y. Gu, G. Zhu, H. Zhou, Q. Gao, R. Huang, J. Cao, and B. Greenhouse. 2020b. “Confirmation of the absence of local transmission and geographic assignment of imported falciparum malaria cases to China using microsatellite panel.” Malar J 19 (1):244. doi: 10.1186/s12936-020-03316-3.

Lo, E., E. Hemming-Schroeder, D. Yewhalaw, J. Nguyen, E. Kebede, E. Zemene, S. Getachew, K. Tushune, D. Zhong, G. Zhou, B. Petros, and G. Yan. 2017. “Transmission dynamics of co-endemic Plasmodium vivax and P. falciparum in Ethiopia and prevalence of antimalarial resistant genotypes.” PLoS Neglected Tropical Diseases 11 (7). doi: 10.1371/journal.pntd.0005806.

Lo, E., N. Lam, E. Hemming-Schroeder, J. Nguyen, G. Zhou, M. C. Lee, Z. Yang, L. Cui, and G. Yan. 2017. “Frequent Spread of Plasmodium vivax Malaria Maintains High Genetic Diversity at the Myanmar-China Border, Without Distance and Landscape Barriers.” J Infect Dis 216 (10):1254–1263. doi: 10.1093/infdis/jix106.

Manrique, P., J. Miranda-Alban, J. Alarcon-Baldeon, R. Ramirez, G. Carrasco-Escobar, H. Herrera, M. Guzman-Guzman, A. Rosas-Aguirre, A. Llanos-Cuentas, J. M. Vinetz, A. A. Escalante, and D. Gamboa. 2019. “Microsatellite analysis reveals connectivity among geographically distant transmission zones of Plasmodium vivax in the Peruvian Amazon: A critical barrier to regional malaria elimination.” PLoS Negl Trop Dis 13 (11):e0007876. doi: 10.1371/journal.pntd.0007876.

Mbunge, Elliot, Richard C Milham, Maureen Nokuthula Sibiya, and Sam Takavarasha Jr. 2023. ”Machine learning techniques for predicting malaria: Unpacking emerging challenges and opportunities for tackling malaria in sub-saharan Africa.” Computer Science On-line Conference.

Mobegi, Victor A, Kovana M Loua, Ambroise D Ahouidi, Judith Satoguina, Davis C Nwakanma, Alfred Amambua-Ngwa, and David J Conway. 2012. “Population genetic structure of Plasmodium falciparum across a region of diverse endemicity in West Africa.” Malaria journal 11:1–9.

Neafsey, Daniel E, Aimee R Taylor, and Bronwyn L MacInnis. 2021. “Advances and opportunities in malaria population genomics.” Nature Reviews Genetics 22 (8):502–517.

Osorio, L., J. Todd, R. Pearce, and D. J. Bradley. 2007. “The role of imported cases in the epidemiology of urban Plasmodium falciparum malaria in Quibdó, Colombia.” Trop Med Int Health 12 (3):331–41. doi: 10.1111/j.1365-3156.2006.01791.x.

Pacheco, M. A., K. A. Schneider, N. Céspedes, S. Herrera, M. Arévalo-Herrera, and A. A. Escalante. 2018. “Limited differentiation among Plasmodium vivax populations from the northwest and to the south Pacific Coast of Colombia: A malaria corridor?” PLoS Neglected Tropical Diseases 13 (3). doi: 10.1371/journal.pntd.0007310.

Patel, J. C., S. M. Taylor, P. C. Juliao, C. M. Parobek, M. Janko, L. D. Gonzalez, L. Ortiz, N. Padilla, A. K. Tshefu, M. Emch, V. Udhayakumar, K. Lindblade, and S. R. Meshnick. 2014. “Genetic Evidence of Importation of Drug-Resistant Plasmodium falciparum to Guatemala from the Democratic Republic of the Congo.” Emerg Infect Dis 20 (6):932–40. doi: 10.3201/eid2006.131204.

Phelan, J. E., A. Turkiewicz, E. Manko, J. Thorpe, L. N. Vanheer, M. van de Vegte-bolmer, N. T. H. Ngoc, N. T. H. Binh, N. Q. Thieu, J. Gitaka, D. Nolder, K. B. Beshir, J. G. Dombrowski, S. M. Di Santi, T. Bousema, C. J. Sutherland, S. Campino, and T. G. Clark. 2023. “Rapid profiling of *Plasmodium* parasites from genome sequences to assist malaria control.” Genome Medicine 15 (1):11. doi: 10.1186/s13073-023-01247-7.

Preston, Mark D, Susana Campino, Samuel A Assefa, Diego F Echeverry, Harold Ocholla, Alfred Amambua-Ngwa, Lindsay B Stewart, David J Conway, Steffen Borrmann, and Pascal Michon. 2014. “A barcode of organellar genome polymorphisms identifies the geographic origin of Plasmodium falciparum strains.” Nature communications 5 (1):4052.

Roh, M. E., S. K. Tessema, M. Murphy, N. Nhlabathi, N. Mkhonta, S. Vilakati, N. Ntshalintshali, M. Saini, G. Maphalala, A. Chen, J. Wilheim, L. Prach, R. Gosling, S. Kunene, S. Hsiang M, and B. Greenhouse. 2019. “High Genetic Diversity of Plasmodium falciparum in the Low-Transmission Setting of the Kingdom of Eswatini.” J Infect Dis 220 (8):1346–1354. doi: 10.1093/infdis/jiz305.

Rosenthal, Philip J. 2022. “Malaria in 2022: challenges and progress.” The American Journal of Tropical Medicine and Hygiene 106 (6):1565.

Schmedes, Sarah E, Dhruviben Patel, Julia Kelley, Venkatachalam Udhayakumar, and Eldin Talundzic. 2019. “Using the Plasmodium mitochondrial genome for classifying mixed-species infections and inferring the geographical origin of P. falciparum parasites imported to the US.” PloS one 14 (4):e0215754.

Sekine, S., C. W. Chan, M. Kalkoa, S. Yamar, H. Iata, G. Taleo, A. Kc, W. Kagaya, Y. Kido, and A. Kaneko. 2024. “Tracing the origins of Plasmodium vivax resurgence after malaria elimination on Aneityum Island in Vanuatu.” Commun Med (Lond) 4 (1):91. doi: 10.1038/s43856-024-00524-9.

Tam, Greta, Benjamin J Cowling, and Richard J Maude. 2021. “Analysing human population movement data for malaria control and elimination.” Malaria Journal 20:1–9.

Tessema, S., A. Wesolowski, A. Chen, M. Murphy, J. Wilheim, A. R. Mupiri, N. W. Ruktanonchai, V. A. Alegana, A. J. Tatem, M. Tambo, B. Didier, J. M. Cohen, A. Bennett, H. J. Sturrock, R. Gosling, M. S. Hsiang, D. L. Smith, D. R. Mumbengegwi, J. L. Smith, and B. Greenhouse. 2019. “Using parasite genetic and human mobility data to infer local and cross-border malaria connectivity in Southern Africa.” Elife 8. doi: 10.7554/eLife.43510.

Trimarsanto, H., R. Amato, R. D. Pearson, E. Sutanto, R. Noviyanti, L. Trianty, J. Marfurt, Z. Pava, D. F. Echeverry, T. M. Lopera-Mesa, L. M. Montenegro, A. Tobón-Castaño, M. J. Grigg, B. Barber, T. William, N. M. Anstey, S. Getachew, B. Petros, A. Aseffa, A. Assefa, A. G. Rahim, N. H. Chau, T. T. Hien, M. S. Alam, W. A. Khan, B. Ley, K. Thriemer, S. Wangchuck, Y. Hamedi, I. Adam, Y. Liu, Q. Gao, K. Sriprawat, M. U. Ferreira, M. Laman, A. Barry, I. Mueller, M. V. G. Lacerda, A. Llanos-Cuentas, S. Krudsood, C. Lon, R. Mohammed, D. Yilma, D. B. Pereira, F. E. J. Espino, C. S. Chu, I. D. Vélez, C. Namaik-larp, M. F. Villegas, J. A. Green, G. Koh, J. C. Rayner, E. Drury, S. Gonçalves, V. Simpson, O. Miotto, A. Miles, N. J. White, F. Nosten, D. P. Kwiatkowski, R. N. Price, and S. Auburn. 2022. “A molecular barcode and web-based data analysis tool to identify imported Plasmodium vivax malaria.” 5 (1). doi: 10.1038/s42003-022-04352-2.

Valdivia, H. O., F. E. Villena, S. E. Lizewski, J. Garcia, J. Alger, and D. K. Bishop. 2020. “Genomic surveillance of Plasmodium falciparum and Plasmodium vivax cases at the University Hospital in Tegucigalpa, Honduras.” Sci Rep 10 (1):20975. doi: 10.1038/s41598-020-78103-w.

Venkatesan, Priya. 2024. “The 2023 WHO World malaria report.” The Lancet Microbe 5 (3):e214.

Ventocilla, J. A., J. Nuñez, L. L. Tapia, C. M. Lucas, S. R. Manock, A. G. Lescano, K. A. Edgel, and P. C. F. Graf. 2018. ”Genetic Variability of Plasmodium vivax in the North Coast of Peru and the Ecuadorian Amazon Basin.” Am J Trop Med Hyg 99 (1):27–32. doi: 10.4269/ajtmh.17-0498.

Wasakul, Varanya, Areeya Disratthakit, Mayfong Mayxay, Keobouphaphone Chindavongsa, Viengphone Sengsavath, Nguyen Thuy-Nhien, Richard D Pearson, Sonexay Phalivong, Saiamphone Xayvanghang, and Richard J Maude. 2023. “Malaria outbreak in Laos driven by a selective sweep for Plasmodium falciparum kelch13 R539T mutants: a genetic epidemiology analysis.” The Lancet Infectious Diseases 23 (5):568–577.

Wesolowski, Amy, Nathan Eagle, Andrew J Tatem, David L Smith, Abdisalan M Noor, Robert W Snow, and Caroline O Buckee. 2012. “Quantifying the impact of human mobility on malaria.” Science 338 (6104):267–270.

Wesolowski, Amy, Gillian Stresman, Nathan Eagle, Jennifer Stevenson, Chrispin Owaga, Elizabeth Marube, Teun Bousema, Christopher Drakeley, Jonathan Cox, and Caroline O Buckee. 2014. “Quantifying travel behavior for infectious disease research: a comparison of data from surveys and mobile phones.” Scientific reports 4 (1):5678.

Wesolowski, Amy, Aimee R Taylor, Hsiao-Han Chang, Robert Verity, Sofonias Tessema, Jeffrey A Bailey, T Alex Perkins, Daniel E Neafsey, Bryan Greenhouse, and Caroline O Buckee. 2018. “Mapping malaria by combining parasite genomic and epidemiologic data.” BMC medicine 16:1–8.

Winter, David J, M Andreína Pacheco, Andres F Vallejo, Rachel S Schwartz, Myriam Arevalo-Herrera, Socrates Herrera, Reed A Cartwright, and Ananias A Escalante. 2015. “Whole genome sequencing of field isolates reveals extensive genetic diversity in Plasmodium vivax from Colombia.” PLoS neglected tropical diseases 9 (12):e0004252.

Xu, S. J., H. M. Shen, Y. B. Cui, S. B. Chen, B. Xu, and J. H. Chen. 2023. “Genetic diversity and natural selection of rif gene (PF3D7_1254800) in the Plasmodium falciparum global populations.” 254. doi: 10.1016/j.molbiopara.2023.111558.

