## Appendix 1: Search commands used in various databases to identify relevant articles for "Detecting imported malaria infections in endemic settings using molecular surveillance: current state and challenges"

**SCOPUS**

TITLE-ABS-KEY ( "Human mobility" OR "Imported case" OR "Imported cases" OR "Imported malaria" OR “Imported Infection” OR “Human migration” OR “Infected individual” OR “Infected individuals” OR “Imported isolate” OR “Imported isolates”) AND TITLE-ABS-KEY (“Connectivity” OR “Gene flow" OR "Genetic surveillance" OR “Genomic surveillance” OR “Transmission”) AND TITLE-ABS-KEY ( "Whole genome sequencing" OR "WGS" OR "Next generation sequencing" OR "NGS" OR "Single nucleotide polymorphism " OR "Single nucleotide polymorphisms" OR "SNP" OR "SNPs" OR "Microsatellite" OR " Sequencing" OR “Genotyping”) AND TITLE-ABS-KEY ( "Malaria" OR "Plasmodium falciparum" OR "Plasmodium vivax" OR "P. falciparum" OR "P. vivax")

TITLE-ABS-KEY ( "Human mobility" OR "Imported case" OR "Imported cases" OR "Imported malaria" OR “Imported Infection” OR “Human migration” OR “Infected individual” OR “Infected individuals” OR “Imported isolate” OR “Imported isolates”) AND TITLE-ABS-KEY ( "Whole genome sequencing" OR "WGS" OR "Next generation sequencing" OR "NGS" OR "Single nucleotide polymorphism " OR "Single nucleotide polymorphisms" OR "SNP" OR "SNPs" OR "Microsatellite" OR " Sequencing" OR “Genotyping”) AND TITLE-ABS-KEY ( "Malaria" OR "Plasmodium falciparum" OR "Plasmodium vivax" OR "P. falciparum" OR "P. vivax")

**PubMed**

(((((((((((Human mobility[Title/Abstract]) OR (Imported case[Title/Abstract])) OR (Imported cases[Title/Abstract])) OR (Imported malaria[Title/Abstract])) OR (Imported Infection[Title/Abstract])) OR (Human migration[Title/Abstract])) OR (Infected individual[Title/Abstract])) OR (Infected individuals[Title/Abstract])) OR (Imported isolate[Title/Abstract])) OR (Imported isolates[Title/Abstract])) AND (((((Connectivity[Title/Abstract]) OR (Gene flow[Title/Abstract])) OR (Genetic surveillance[Title/Abstract])) OR (Genomic surveillance[Title/Abstract])) OR (Transmission[Title/Abstract]))) AND ((((((((((Whole genome sequencing[Title/Abstract]) OR (WGS[Title/Abstract])) OR (Next generation sequencing[Title/Abstract])) OR (NGS[Title/Abstract])) OR (Single nucleotide polymorphism[Title/Abstract])) OR (Single nucleotide polymorphisms[Title/Abstract])) OR (SNP[Title/Abstract])) OR (SNPs[Title/Abstract])) OR (Microsatellite[Title/Abstract])) OR (Sequencing[Title/Abstract])) OR (Genotyping[Title/Abstract]))) AND (((((Malaria[Title/Abstract]) OR (Plasmodium falciparum[Title/Abstract])) OR (Plasmodium vivax[Title/Abstract])) OR (P. falciparum[Title/Abstract])) OR (P. vivax[Title/Abstract]))

(((((((((((Human mobility[Text Word]) OR (Imported case[Text Word])) OR (Imported cases[Text Word])) OR (Imported malaria[Text Word])) OR (Imported Infection[Text Word])) OR (Human migration[Text Word])) OR (Infected individual[Text Word])) OR (Infected individuals[Text Word])) OR (Imported isolate[Text Word])) OR (Imported isolates[Text Word])) AND (((((Connectivity[Text Word]) OR (Gene flow[Text Word])) OR (Genetic surveillance[Text Word])) OR (Genomic surveillance[Text Word])) OR (Transmission[Text Word]))) AND ((((((((((Whole genome sequencing[Text Word]) OR (WGS[Text Word])) OR (Next generation sequencing[Text Word])) OR (NGS[Text Word])) OR (Single nucleotide polymorphism[Text Word])) OR (Single nucleotide polymorphisms[Text Word])) OR (SNP[Text Word])) OR (SNPs[Text Word])) OR (Microsatellite[Text Word])) OR (Sequencing[Text Word])) OR (Genotyping[Text Word]))) AND (((((Malaria[Text Word]) OR (Plasmodium falciparum[Text Word])) OR (Plasmodium vivax[Text Word])) OR (P. falciparum[Text Word])) OR (P. vivax[Text Word]))

((((((((((Human mobility[Title/Abstract]) OR (Imported case[Title/Abstract])) OR (Imported cases[Title/Abstract])) OR (Imported malaria[Title/Abstract])) OR (Imported Infection[Title/Abstract])) OR (Human migration[Title/Abstract])) OR (Infected individual[Title/Abstract])) OR (Infected individuals[Title/Abstract])) OR (Imported isolate[Title/Abstract])) OR (Imported isolates[Title/Abstract])) AND (((((((((((Whole genome sequencing[Title/Abstract]) OR (WGS[Title/Abstract])) OR (Next generation sequencing[Title/Abstract])) OR (NGS[Title/Abstract])) OR (Single nucleotide polymorphism[Title/Abstract])) OR (Single nucleotide polymorphisms[Title/Abstract])) OR (SNP[Title/Abstract])) OR (SNPs[Title/Abstract])) OR (Microsatellite[Title/Abstract])) OR (Sequencing[Title/Abstract])) OR (Genotyping[Title/Abstract]))) AND (((((Malaria[Title/Abstract]) OR (Plasmodium falciparum[Title/Abstract])) OR (Plasmodium vivax[Title/Abstract])) OR (P. falciparum[Title/Abstract])) OR (P. vivax[Title/Abstract]))

((((((((((Human mobility[Text Word]) OR (Imported case[Text Word])) OR (Imported cases[Text Word])) OR (Imported malaria[Text Word])) OR (Imported Infection[Text Word])) OR (Human migration[Text Word])) OR (Infected individual[Text Word])) OR (Infected individuals[Text Word])) OR (Imported isolate[Text Word])) OR (Imported isolates[Text Word])) AND (((((((((((Whole genome sequencing[Text Word]) OR (WGS[Text Word])) OR (Next generation sequencing[Text Word])) OR (NGS[Text Word])) OR (Single nucleotide polymorphism[Text Word])) OR (Single nucleotide polymorphisms[Text Word])) OR (SNP[Text Word])) OR (SNPs[Text Word])) OR (Microsatellite[Text Word])) OR (Sequencing[Text Word])) OR (Genotyping[Text Word]))) AND (((((Malaria[Text Word]) OR (Plasmodium falciparum[Text Word])) OR (Plasmodium vivax[Text Word])) OR (P. falciparum[Text Word])) OR (P. vivax[Text Word]))

**Cochrane Library**

("Human mobility" OR "Imported case" OR "Imported cases" OR "Imported malaria"  OR “Imported infection” OR "Human migration" OR “Infected individual” OR “Infected individuals” OR “Imported isolate” OR “Imported isolates”) AND (("Connectivity" OR "Gene flow" OR "Genetic surveillance" OR "Genomic surveillance" OR "Transmission") AND ("Whole genome sequencing" OR "WGS" OR "Next generation sequencing" OR "NGS" OR "Single nucleotide polymorphism" OR "Single nucleotide polymorphisms" OR "SNP" OR "SNPs" OR "Microsatellite" OR "Sequencing" OR "Genotyping")) AND (("Malaria" OR "Plasmodium falciparum" OR "Plasmodium vivax" OR "P. falciparum" OR "P. vivax"))

("Human mobility" OR "Imported case" OR "Imported cases" OR "Imported malaria"  OR “Imported infection” OR "Human migration" OR “Infected individual” OR “Infected individuals” OR “Imported isolate” OR “Imported isolates”) AND (("Whole genome sequencing" OR "WGS" OR "Next generation sequencing" OR "NGS" OR "Single nucleotide polymorphism" OR "Single nucleotide polymorphisms" OR "SNP" OR "SNPs" OR "Microsatellite" OR "Sequencing" OR "Genotyping")) AND (("Malaria" OR "Plasmodium falciparum" OR "Plasmodium vivax" OR "P. falciparum" OR "P. vivax"))

**Web of Science**

((((((((AB=(Human mobility)) OR AB=(Imported case)) OR AB=(Imported cases)) OR AB=(Imported malaria)) OR AB=(Imported Infection)) OR AB=(Human migration)) OR AB=(Infected individual)) OR AB=(Infected individuals)) OR AB=(Imported isolate)) OR AB=(Imported isolates) AND ((((AB=(Connectivity)) OR AB=(Gene flow)) OR AB=(Genetic surveillance)) OR AB=(Genomic surveillance)) OR AB=(Transmission) AND ((((((((((AB=(Whole genome sequencing)) OR AB=(WGS)) OR AB=(Next generation sequencing)) OR AB=(NGS)) OR AB=(Single nucleotide polymorphism )) OR AB=(Single nucleotide polymorphisms)) OR AB=(SNP)) OR AB=(SNPs)) OR AB=(Microsatellite)) OR AB=(Sequencing)) OR AB=(Genotyping) AND ((((AB=(Malaria)) OR AB=(Plasmodium falciparum)) OR AB=(Plasmodium vivax)) OR AB=(P. falciparum)) OR AB=(P. vivax)
